# Modeling vaccine allocation and equity implications of COVID-19 containment strategies

**DOI:** 10.1101/2022.09.05.22279623

**Authors:** Ichiro Nakamoto

## Abstract

Given the shortage of global COVID-19 vaccines, a critical public concern is whether the strategy of allocation exerts a heterogeneous effect on settings that have imbalanced accessibility. Exacerbated by the mutational characteristics of the pathogen, traits of immunity protection of vaccines, and diversification of human behaviors, the pathway to the full eradication of the COVID-19 pandemic is becoming increasingly complicated and indeterminate. Population-wide evaluation of public interventions remains crucial to evaluate the performance of epidemiology policies. This study employs a mathematical compartmental model combined with the observational data of the United States to examine the potential effect of vaccine allocation on the trajectory of COVID-19 transmission and the elicited equity implications. The outcomes imply that allocation strategies substantially impact the cumulative equilibrium size of a pandemic controlling for confounding factors. Under a framework of a two-dose primary vaccination strategy aiming to curb the total infections for high-accessibility settings (HAS) and low-accessibility settings(LAS), the traits of vaccination, pathogen, and human effort integrally affect the equilibrium of the COVID-19 pandemic in the medium perspective (i.e., up to 5 years). Vaccine allocation increases the healthcare and cost burden for HAS temporarily, in contrast, it reduces the risk of COVID-19 transmission for the LAS. The effects are consistent across a variety of profiles. By enhancing the administration rates of primary doses (i.e., mainly through dose 1 and dose 2), the magnitude of the COVID-19 pandemic decreases contingent on confounding factors. To minimize the magnitude of infection, it is of importance to dynamically monitor the immunity protection of vaccines, the dynamics of virus transmission, and the gap in the human effort.

---

As of June 2022, globally more than 0.5 billion cases of SARS-CoV-2 infections were officially identified according to the WHO surveillance data ^1^. Studies have uncovered evidence that the strategies of vaccine distribution exert crucial roles in influencing the trajectory of COVID-19 transmission ^2,3,4,5^. Equitable and efficient provision of affordable vaccines is projected to be positively connected with the mitigation and depletion of a pandemic ^6^. Re-allocation of vaccines across places that have a gap in accessibility to medical resources provides a potential channel to optimize the integral benefits of vaccines ^7,8^. The Vaccines Global Access Facility (COVAX) framework supported by WHO aims to yield equitable vaccine supplies, which is a procurement proposal leveraging the imbalance between well-served and under-served communities ^9,10^. Nevertheless, attributable to elaborate factors such as sub-optimal supports, COVAX has difficulty in meeting its original targets ^9,10,11,12,13^. The inequality of vaccine distribution raises important challenges, which can significantly erode the collaborative efforts of many countries and potentially extend the global timeline of the COVID-19 epidemic^14,15^.

One of the major concerns of vaccine allocation is to optimize the output of public health as a whole. However, rigorous quantifying the optimal strategy of allocation is challenging because it could be influenced by perplexing identifiable and non-identifiable factors ^16^. On the other hand, research shows that the flexibility of allocation strategies can enhance the efficiency, and reduce severe outcomes and the operation costs of the healthcare system when resource provision is constrained ^17,18^. With the decline of immunity protection over time, the risks of exposure to infections could worsen ^19,20^. The recent emergence of novel variants of COVID-19 identified in low- and middle-income countries indicates that low accessibility to vaccines could remarkably deteriorate the transmission of COVID-19 ^21^. Effective vaccines can enhance the immunity protection for individuals and hence lower the risks of being infected at exposure ^22,23,24^. Studies have confirmed that vaccination could facilitate the formation of a decent level of protection in the absence of severe variants ^25,26^, on the other hand, the waning of immunity increases the risks of reinfection, which triggers the necessity of social distancing and subsequent vaccination ^27,28,29,30^.

The topic of vaccine allocation is contentious. Debates remain on the potential pros and cons incurred ^31,32,33,34^. To explore the impact of vaccine allocation and the principle mechanism that affects the transmission dynamics of the pandemic, we employ a two-dose mathematical compartmental model targeting two settings that are not balanced in vaccine accessibility ^3^. We evaluate the medium-term effects up to 5 years of vaccine allocation coupled with the traits of the vaccines, characteristics of the pathogen, and efforts of human behaviors. We aim to investigate (1) whether vaccine allocation could affect the equilibrium of disease infections across settings; (2) how and to what extent the effects change contingent on the level of mutations; (3) how traits of the vaccines, characteristics of the pathogen, and efforts of human behaviors influence the transmission respectively; (4) whether the ratio of allocation affects the trajectory of transmission. To mimic the various scenarios in practice, we additionally assume that (1) HAS and LAS are not equal in the accessibility to vaccines; (2) individuals are free to move between settings (Fig. 1).

**Fig. 1.**
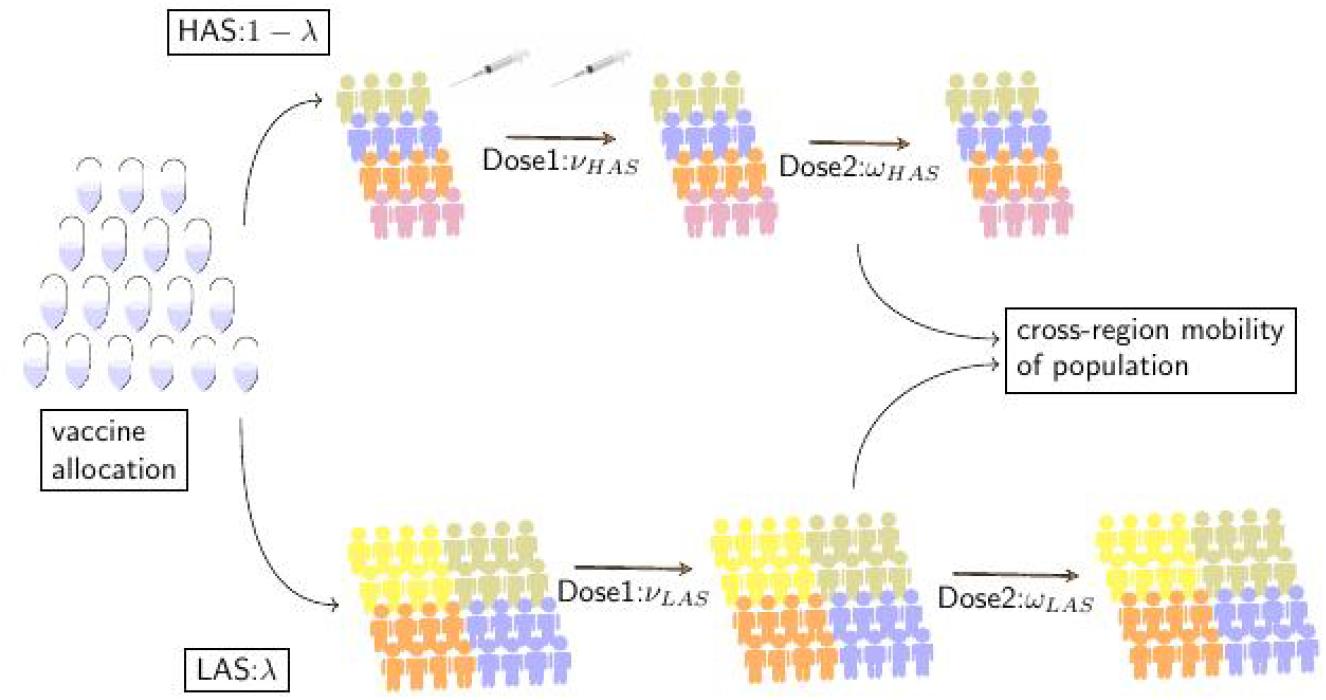
COVID-19 vaccine allocation and flowchart of transmission dynamics. Assume the populations are divided into two settings: the first group is defined as the high-accessibility setting(HAS) and the second group is termed as low-accessibility setting(LAS). The gap in vaccine accessibility is present across the two settings. A two-dose vaccination program is scheduled to impart partial immunity protection to the population Individuals are not restricted to move across the settings Individuals vaccinated with dose 1 or dose 2 lost immunity protection over time and are exposed to the risks of secondary infection. The strength of pathogen transmission across the two settings is subjective to a variety of profiles.

Our results show that to optimize the health output of the public, vaccine allocation can reduce the total cumulative infections of settings at equilibrium in the absence of severe variants. The optimal ratio of allocation is correlated with transmission rates, rollout speeds, immunity protection, rate of waning, rate of birth, rate of death, rate of recovery, and risks of secondary infection. Efficient distribution of primary doses, which is mainly through one-dose or two-dose series, is critical to mitigate or even deplete the scale of the epidemic^22^. The outcomes are broadly consistent across a variety of scenarios.

## Results

### Vaccine allocation reduces the total equilibrium incidence of COVID-19 infections from the medium perspective

To evaluate the effect of vaccine allocation between two settings that have unbalanced accessibility to vaccine resources on the transmission dynamics of COVID-19, we performed simulation under a variety of profiles including no population mobility, low population mobility, medium population mobility, and high population mobility respectively (Fig. 2A-2H). The assumptions are (a) the initial scale of infection is identical across the two settings; (b) the rate of birth is equal to the rate of death;(c) the timing of vaccination and vaccine allocation from HAS to LAS is initiated at week 16, nearly fourth months after the pathogen transmission is established; (d) After reallocation, rollout rate for HAS is decreased, in contrast, the rate for LAS is increased attributable to the change in availability; (e) dose 2 confers better protection than dose 1 and the immunity protection of vaccines wanes over time.

**Fig 2.**
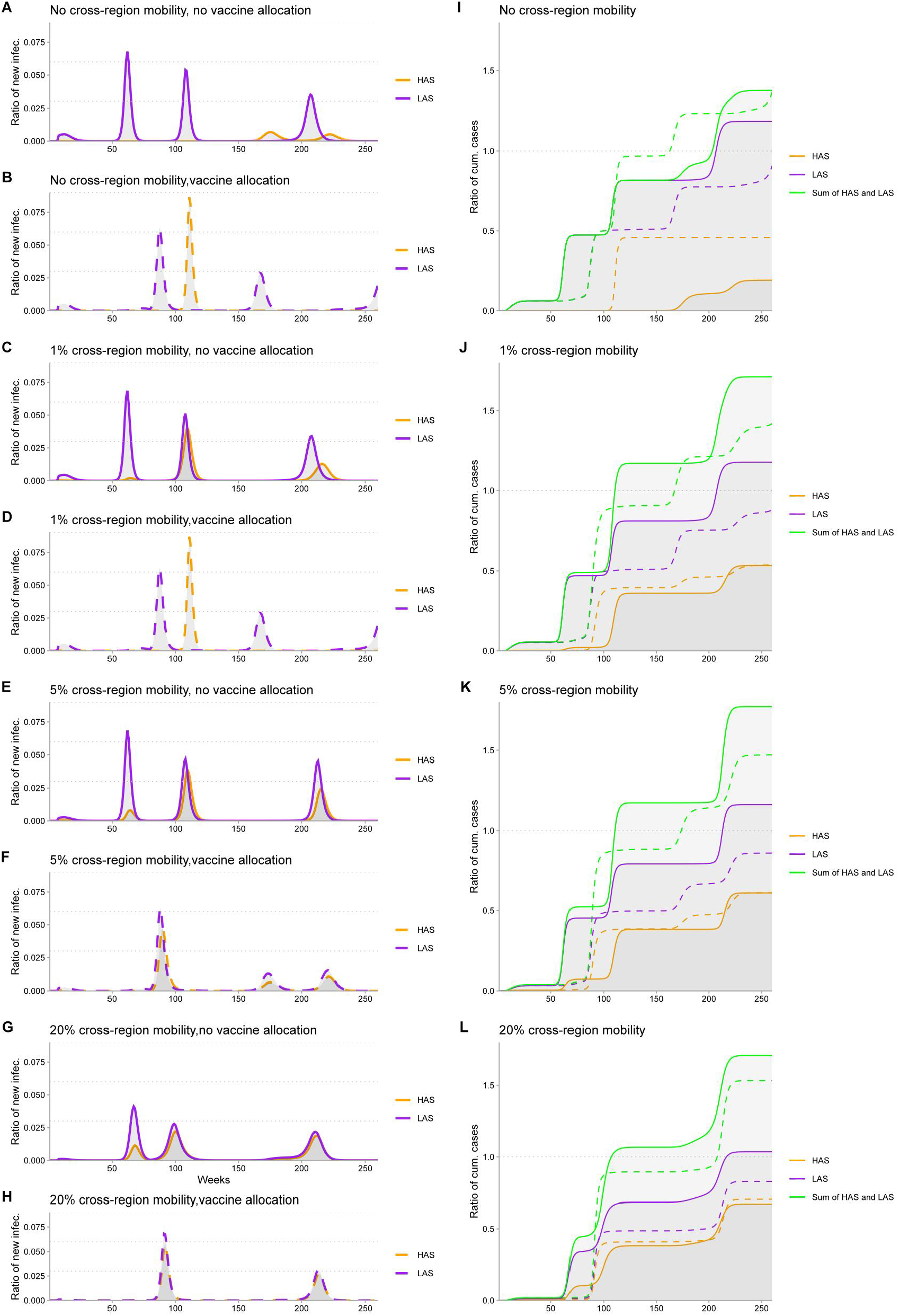
Impact of vaccine allocation and population mobility on the transmission dynamics of COVlO-19 pandemic Allocation ratio of vaccines from HAS to LAS is 0 5

In all scenarios, reallocation of vaccines from HAS to LAS reduces the total equilibrium cases of COVID-19 infections, which is consistent across differentiated mobility profiles. Generally, reallocation of vaccines deteriorates the circumstance of infection in HAS, in contrast, it improves the circumstance in LAS from a medium perspective for up to five years. At the initial stage of allocation, the overall burden aggregating HAS and LAS is temporarily enlarged, however, with the advancement of vaccine redistribution, the integrated curtailing effect does not change qualitatively. The magnitude of COVID-19 infection is relatively smaller for the no-mobility scenario in comparison to other mobility scenarios. Asymptotically, the ultimate scale of infection at equilibrium remains at a commensurate level for the mobility profiles, in contrast, the value is reduced from around 1.7 percent to 1.4 percent, nearly 0.3 point of reduction in magnitude (Fig. 2I versus J, K, and L). We evaluate the case where ratio of allocation equal to 50%, other scenarios of vaccine allocation including 10%, 30%, 70% and 90% are illustrated in the supplementary document (Fig. S2-S5, Table S1) and the outcomes are qualitatively identical viewing from a medium-term perspective.

### The efforts of human behavior (i.e., vaccination rates) substantially impact the trajectory of COVID-19 in the absence of severe variants

To analyze how human behavior impacts the transmission dynamic, we performed simulations by employing differentiated vaccination rates to mimic a variety of delineations where rollout inefficiency (*ν* = 0.0001), normal vaccination (*ν* = 0.003), and speed-enhanced vaccination of dose 1 are observed respectively (*ν*= 0.005, Fig. 3 A-F). In a similar vein, we compare two scenarios regarding dose 2 (*ω* = 0.02 versus *ω*= 0.05). We subsequently evaluate how administration rates, immunity protection, immunity waning, infection risks, transmission risks, rate of birth, rate of death, recovery rate, and their interactions exert influence on the cumulative equilibrium status across the two imbalanced settings. We assume three differentiated levels of exposure to secondary infection (*ε* = 0.5, 0.6, 0.7) to emulate the potential risks that individuals encounter after the immunity protection wanes. To account for prospective diversified scenarios of transmission in practice, we additionally incorporate three cases where: (1) transmission risk in HAS and LAS is at commensurate levels; (2)transmission risk of HAS is more severe; (3)transmission risk of LAS is more severe. All other parameters utilized in the analysis are illustrated in detail (See Supplementary Table S2).

**Fig. 3.**
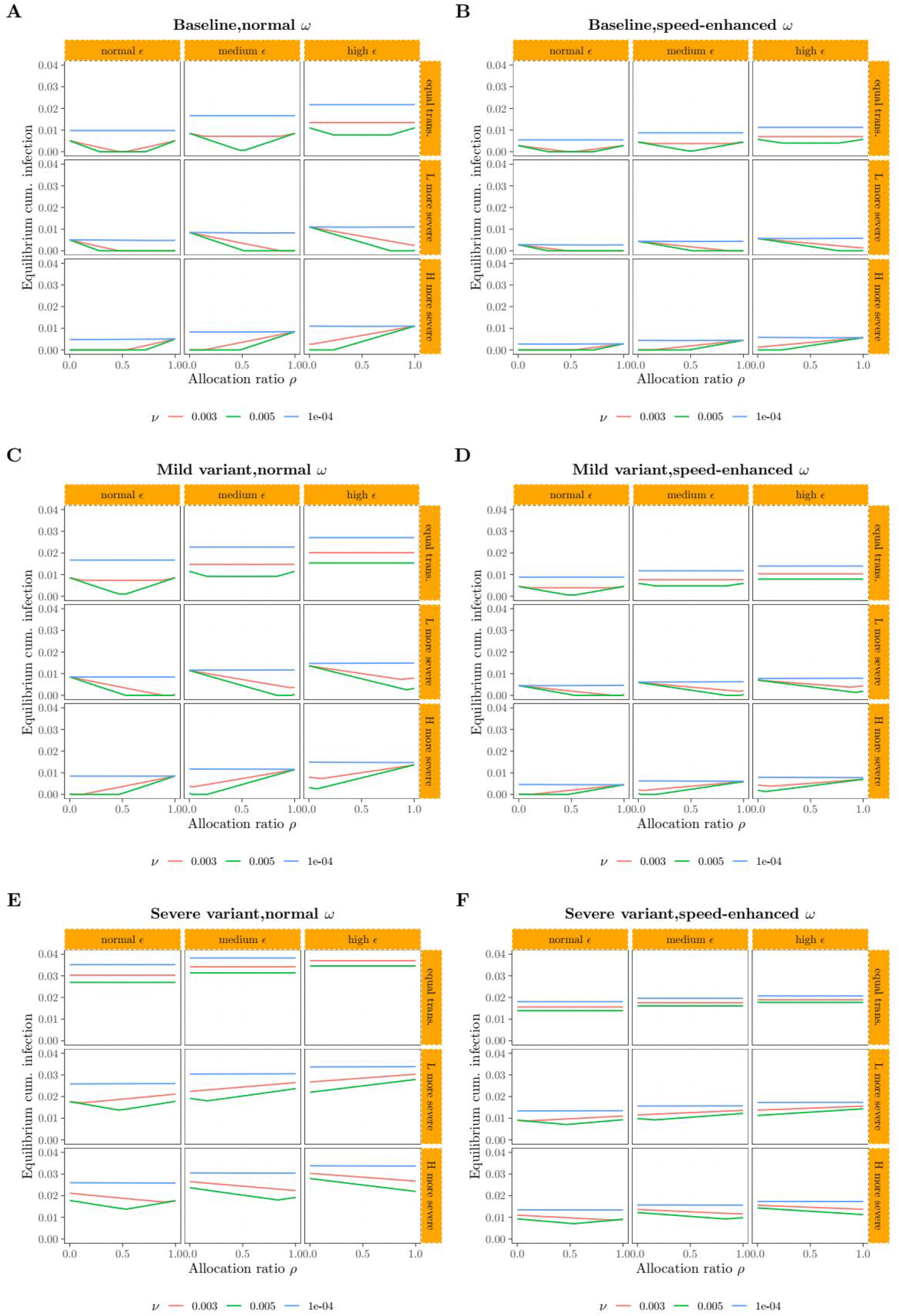
Impact of vaccination rate on the COVID-19 transmission.

The trajectory of the COVID-19 pandemic is not affected noticeably if the rates of vaccination, either dose 1 or dose 2, are inefficient (i.e, close to boundary) under the framework of vaccine allocation (Fig. 3 A-F panel blue curves). To minimize the size of cumulative equilibrium infection, the optimal allocation ratio varies contingent on the rate of vaccination, risk levels of transmission, and risk of secondary infection. The rate of vaccination plays a critical role in altering the advancement of the epidemic. Speedier rollout of dose 1 or dose 2 reduces the magnitude of cumulative equilibrium cases of COVID-19 ceteris paribus(Fig. 3 A versus B, C versus D, E versus F). In the case where the magnitude of transmission risk for HAS is at the commensurate level relative to LAS, the total number of equilibrium infections follows a regular trend with the growth of allocation ratio. Initially the infection decreases monotonically and diminishes to zero when the allocation ratio reaches a threshold. After this point, the growth of allocation does not change the status of equilibrium. However, the disease-free equilibrium starts to break and the number of cases increases when the allocation ratio continues and exceeds a threshold point where the effect of allocation is sub-optimal (Fig. 3, red curve in upper panel). When LAS is exposed to higher transmission risks, vaccine allocation from HAS to LAS consistently curtails the transmission of the pandemic (Fig.3, middle panel). In contrast, the opposite trend is observed when the infection at HAS is more severe (Fig.3, lower panel). In the case where a variant of the pathogen is emerging (Fig. 3C and Fig. 3E), the magnitude of cumulative cases is enlarged ceteris paribus. Generally, severe variants of COVID-19 compromise the protection of vaccines and human efforts (Fig. 3E and Fig. 3F). Hence, the appropriate ratio of vaccine allocation is a crucial factor in influencing the total incidence across settings. Asymptotically, in the baseline scenario where vaccinate rates *ω* = 0.02 and *ν* = 0.003, when the risk of transmission is identical for HAS and LAS, reduction of secondary infection risk from 0.5 to 0.6 increases the minimum scale of equilibrium infection by around 0.6%, and this value changes to nearly 0.8% when the risk grows from 0.6 to 0.7. In contrast, when the rollout rate rises to *ω*= 0.05 and the efficiency is improved, the curtailing effect promotes to approximately 0.4% and 0.3% respectively. Efficiency improvement in the human effort is positively connected to the scale-down of the pandemic.

Multiple factors including the traits of vaccines, the characteristics of pathogen transmission, and the uncertainty of human behavior play roles in impacting the effects of vaccine allocation. If mutations emerge or the risk of infection is increasing, it is of difficulty to contain the spread of the pathogen through the channel of vaccine allocation (Fig. 3A versus C, B versus D). The effect of allocation does not hold when severe variants occur and vaccines lose the strength of protection (Fig. 3E and F). In this case, increasing the rate of rollout slightly changes the trajectory of transmission, however, the effect is generally limited and uncertain.

### Immunity protection conferred by vaccines substantially impacts the equilibrium of COVID-19 infections in the absence of severe variants

We subsequently analyze how the rate of immunity waning, risk of secondary infection, rates of vaccination, and their interactions affect the trajectory of the COVID-19 pandemic. We compare three scenarios where the immunity conferred by vaccines wanes at low, medium, and high speed respectively (Fig. 4A, C, E, and Fig. 4B, D, F). We evaluate three levels of risks of secondary infection (low,medium,high) to mimic similar scenarios after the immunity protection conferred by vaccines wanes. As the quality of healthcare for HAS and LAS could be heterogeneous, we estimate three types of rate ratio 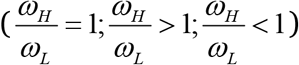 to incorporate the potential impact.

**Fig. 4.**
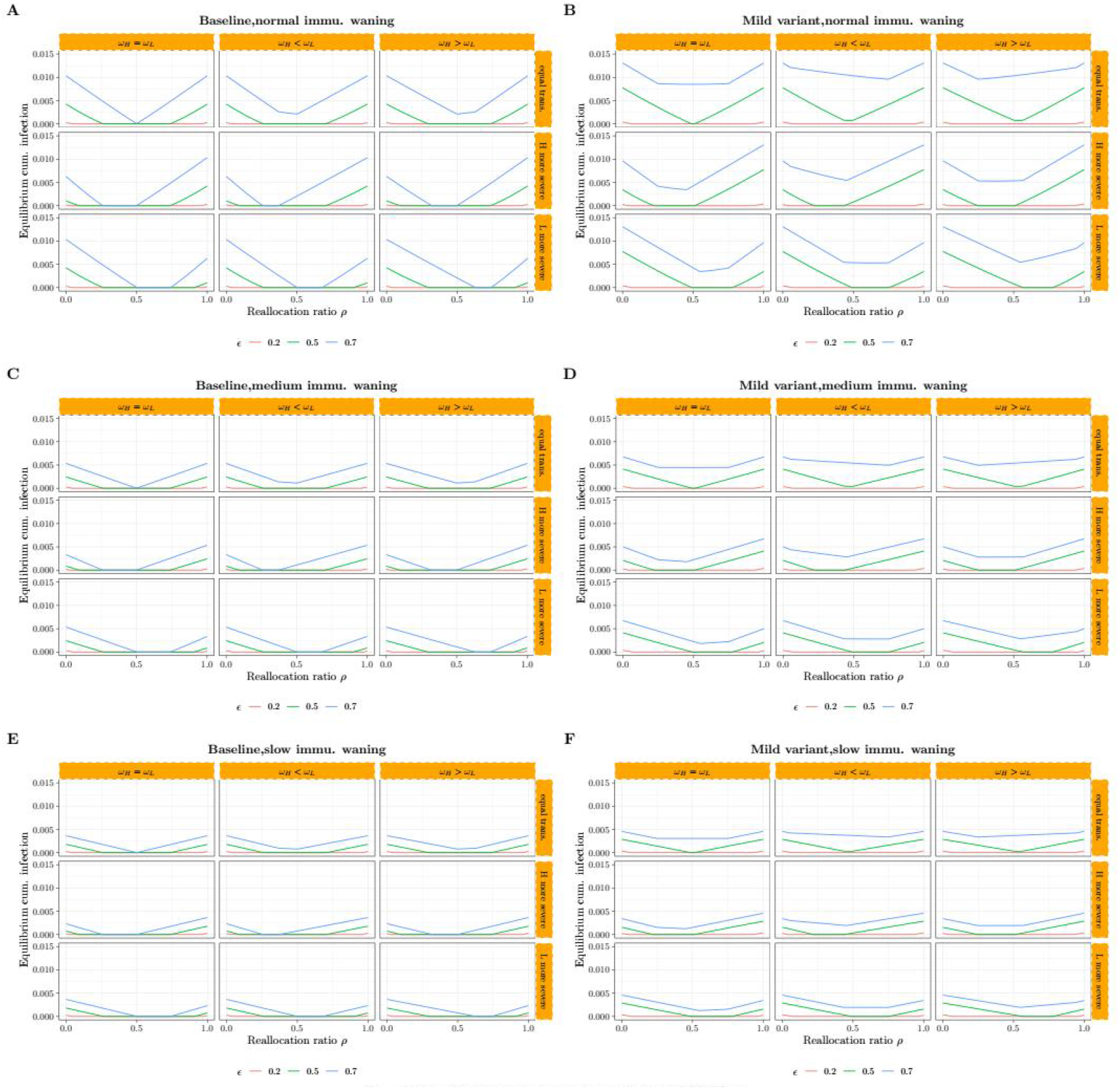
Impact of vaccine immunity on the transmission of COVID-19.

A higher likelihood of secondary infection deteriorates the magnitude of cumulative equilibrium infections (Fig. 4A, blue curve versus green curve versus red curve). In the case where the risk of secondary infection is low or close to the boundary, the effect of vaccine allocation is negligible (Fig. 4A-F, red curves). Consistently, there exists an optimal ratio of vaccine allocation minimizing the magnitude of total equilibrium cases across all profiles controlling for other confounding factors(Fig. 4A-F). After the optimal ratio is passed, growth in vaccine allocation changes the disease-free equilibrium and the number of cases increases subsequently. When the transmission in HAS is riskier, allocation of vaccines enlarges the scale of integral infections if the allocation ratio continuously grows from zero to one; in contrast, the reverse is observed when transmission in LAS is more severe (Fig. 4A middle panel versus lower panel). Stronger immunity protection conferred by vaccines reduces the equilibrium size of the pandemic (Fig. 4A versus 4C versus 4E). Asymptotically, when the risk of transmission for HAS and LAS is identical and high, symmetric pace of rollout is more likely to generate disease-free equilibrium status than otherwise (Fig. 4A, C, blue curves in upper panel).

## Discussion

Our findings suggest that vaccine allocation from HAS to LAS mitigates the burden of pathogen spread and curtails the size of cumulative equilibrium cases. Second, rates of rollout substantially impact the trajectory of the COVID-19 epidemic. Third, we show the asymptotic relation between the three variables including viral, vaccinal, and human factors and the magnitude of infections through mathematical modeling and analysis. Fourth, the effect is sub-optimal in the case where the rollout rate of vaccines is close to the boundary or negligible. Increasing the administration rate of primary dose series can lower the magnitude of total incidence. The findings are robust to diversified risks of transmission, rates of recovery, rates of birth, rates of death, immunity protection, and risks of secondary infection across a variety of profiles. Fifth, the efficiency of vaccination plays a critical role in curbing the size of a pandemic. Hence, how to deploy vaccines effectively and timely is the challenge policymakers need to deliberate under the allocation framework. Although temporarily it induces a burden for HAS, the total benefits of allocation outweigh the costs as a whole.

We explored the correlation between rates of vaccination, rates of transmission, rates of birth/death, rates of immunity waning, and rates of secondary infection in determining the equilibrium scale of the pandemic and thereafter potential improvement of strategy regarding vaccine allocation. In contrast, the effect is negligible when administration capacity is close to boundary-binding (Fig. 4 and Fig. 5). Studies have broad consensus on the general effectiveness of COVID-19 vaccines ^35,36,37,38^. Prior work suggests that immunity protection by vaccines is expected to trigger a sizable effect on curtailing average infections if immunity protection wanes over time after vaccination ^37,38^. The rates at which the COVID-19 vaccines are distributed to vulnerable individuals are critically important and are associated with later shrinking outbreaks ^39,40,41,42,43,44^, we extend prior findings to the vaccine allocation case where vaccinal, pathogenic, and human behavior factors are simultaneously interconnected. The outcomes are sensitive to the severity level of COVID-19.

Studies have identified that fewer than one-half of donated vaccines were administered in some settings, hence the benefits of improving the efficiency of the rollout are multi-fold. Challenges including misinformation, vaccine hesitancy, and compromised public confidence need to be addressed carefully ^10,28,45,46^. Data have documented that the hesitancy rate of the COVID-19 vaccine was more than 20% in some countries ^36,47,48^. Addressing the issue of global vaccine hesitancy is a dilemma that the world needs to take in account carefully ^49,50^.

Debates on the necessity of vaccine allocation remained since the transmission was identified. The allocation of vaccines is motivated by concerns about equal access to medical resources and reduced risk of novel variants globally. In settings where the provision of doses is significantly challenged, donations from their counterparts would render greater pros than cons. A partial burden might be provoked for the donating side, however, it will not outperform the total benefits viewing from a longer perspective. The COVAX program called for assistance to distribute more available vaccines to more populations globally ^29^. New waves of variants were exacerbating the public health crisis worldwide ^51^. The decision-making is elaborate and requires both professional and social welfare considerations, reflecting strategic priorities at specific timing and specific contexts. Decisions for optimal allocation strategies need to be evidence-based and consider the benefits and risks for both local communities and remote counterparts as a whole ^52^, understanding of vaccinal and pathogenic attributes, as well as the efforts delivered by human beings ^53,54,55^.

We investigated the equity implications for the allocation of limited supplies of COVID-19 vaccine doses from HAS to LAS in curtailing the equilibrium size of the pandemic. We examined the level of the equity through the channel of allocation ratio, which indicates that an appropriate ratio of allocation contributes to minimize the cumulative cases of infections and hence generates optimal benefits accounting for transmission risks, rates of rollout, rates of recovery, immunity protection, rates of birth/death and risks of secondary infection. Our findings suggest that rapid deployment and equitable allocation of vaccines to less served settings are preferable if optimization of the public health is the principal objective ^56,57^.

The provision of vaccines is increasing over time but is not uniformly distributed ^15^. Vaccines facilitated in shaping the trajectories of the epidemic in past outbreaks ^58,59,60^. Priorities need to be given to the coverage of vaccine series to enhance the balance level between settings currently as more variants are identified frequently. More observational data will be needed to understand the potential impact of vaccine allocation, particularly in the context of Omicron-like novel variants. In the global context of supply constraints and imbalance, the allocation strategies of doses potentially change the random spread of the pathogen and even deplete the transmission of a pandemic ^59,61^. It is therefore imperative to quantify the immunity protection and immunity waning of vaccines, the biological attributes of the virus through more clinical tests, and collect in-practice data regarding the effort of human behavior to update the allocation policies ^35^.

Our study has several limitations. We do not take into account the stochastic dynamics of the transmission and most of the outcomes are based on SVIR deterministic models. Although we consider the heterogeneity of population mobility, rate of vaccination, and immunity protection, we do not account for other risk factors that are potentially connected with COVID-19 infection including age, inter-person contact, and socioeconomic status. Further stratified analysis may lead to a more refined picture regarding the prioritization sequence and the optimal ratio of allocation for sub-groups. Study found that it could be preferable to distribute doses with priority to immunity-compromised individuals and essential healthcare workers whose risk of being infected is high ^56^. In addition, parameters and information utilized in the model are subject to uncertainty and variation, and the generalizability of the model results needs to be calibrated with the availability of new inputs. For instance, we assume a two-dose framework, however, countries might employ different strategies (i.e, booster doses) and other types of vaccines that are biologically disparate from our assumptions. The optimal strategy of vaccine allocation relies on the trajectory of the epidemic that is specific to the setting, such as reproductive number, contact matrix of population, dynamic immunity protection, and dynamic infectiousness of the pathogen. These features are context specific and uncertain, and the model relies on more observational data. Further, we do not explicitly rationalize the heterogeneity of vaccine hesitancy across settings, which could potentially alter the effect of the vaccine allocation strategy. Evidence is accumulating to update favorable global recommendations of allocation strategy, which may be refined as more data are available. Finally, we assume a single vaccine framework, which might be distinctive from the real scenario ^62,63,64^. As more vaccines are available, decision makers have more options to choose conditional on the heterogeneity inherent in the immunity protection and waning profiles of vaccines, the dynamic of pathogen transmission, and the effort of human behavior. And more clinical data are needed to decide whether allocation strategies assimilating mixing types of vaccines could produce better benefits.

The difficulty in complete containment of the COVID-19 pandemic reinforces the importance of equitable access to COVID-19 vaccines ^64^. Promoting the protection against COVID-19 needs to solve multifaceted challenges including limited vaccine supplies, vaccine hesitancy, and logistical problems, which requires a global perspective on coordination and collaboration. Allocation strategies of COVID-19 vaccines should have clarified objectives and update dynamically with the supply change. Rationing processes need to account for fairness and be subjected to public scrutiny ^65^. Currently, some counties opt to deviate away from COVAX programs ^66,67,68,69^, which reflects another significant challenge ahead. One countermeasure to this issue is to transfer vaccine-relative technologies to LAS-based manufacturers in the future to enhance equity and solidarity ^14^. Vaccine reallocation might not necessarily be the sole solution to solve the issue of imbalance, insufficiency, and inequality ^67^, however, it promotes the level of accessibility for the individuals in under-served settings ^68^.

Most of the new lineages confirmed in low-income settings mostly have mutations that have not been identified elsewhere ^70^. Variants of COVID-19 including Omicron reflected the novel dilemma and the latent risk ahead ^53^. We recommend that allocation frameworks for determining eligibility for efficacious COVID-19 vaccines in the first instance should be evaluated based on viral, vaccinal, and human factors, with the caveat that this might change as vaccine supplies update globally. Consistent criteria that can be subject to objective scrutiny to ensure accountability, equity, and fairness are required ^71^. Various human efforts are of necessity to maintain healthcare operations ^72,73^.

Our model-based evaluation highlights the merits of vaccine allocation across settings that have gaps in vaccine accessibility. The outcomes presented in this study afford insights to asymptotically yield improved allocation strategies contingent on the availability of vaccines, stratified healthcare objectives, dynamics of transmission, human behavior effort, public confidence, vaccine hesitancy, and other potential factors ^62,63^. The optimal vaccine allocation strategy depends on complicated factors including the attributes of pathogen, the traits of vaccines, and the effort of human behaviors. Evaluation of the value of COVID-19 vaccine allocation strategies contributes to the promotion of preparedness and enhances the quality of logistical management of vaccines for future pandemics.

## Supporting information

none

## Data Availability

All data produced in the present study are available upon reasonable request to the authors

## Declarations

### Ethical Approval and Consent to participate

All authors consent to participate.

### Consent for publication

All authors consent for publication.

### Availability of supporting data

No datasets were utilized in this modeling study. Parameter calibrations for simulations were partially obtained from the literature and assumptions as described in Supplementary Table S1-S5. All simulations and analyses were performed using R4.0.3, deSolve package1.28, and dplyr package 1.0.6.

### Authors’ contributions

I.N. conceptualized the study. I.N. designed models and implemented simulations, performed analyses, interpreted results and wrote and revised the manuscript. J.L.Z. and I.N. reviewed and contributed to the revision of the manuscript.

## Acknowledgments

Not applicable.

