## Supplementary material for "Modeling vaccine allocation and equity implications of COVID-19 containment strategies": none

This supplementary text outlines the framework of mathematical models, the flowchart of COVID-19 transmission, equilibrium solutions of the dynamics, and the correlation between the equilibrium, characteristics of the pathogen, traits of immunity protection of vaccines, and diversification of human behaviors. We incorporate the outcomes for a variety level of allocation ratio and population mobility corresponding to the major outcomes of the manuscript. The definitions and values of parameters utilized in the simulations are illustrated subsequently. The supplementary material mainly consists of three sections as follows.

Supplementary Methods
Supplementary Figures S1-S5
Supplementary Table S1-S3

### Supplementary Methods

#### Methods

**Compartmental models.** The mathematical model and assumptions used in this study share the principal immuno-epidemiological mechanism of Fig. 1 and Fig. S1 , incorporating characteristics of the pathogen, traits of immunity protection of vaccines, and diversification of human behaviors. The compartment $S_P$ denotes full susceptible individuals; $S_S$ stands for partially susceptible individuals at exposure to the risk of reinfection. $I_P$ , and $I_S$ represent primary and secondary infections respectively from the corresponding susceptibility compartment; $V_i$ includes individuals vaccinated with dose $i$ ; and compartment $R$ means recovery from the infection. To allow for heterogeneity in rate of vaccination, immune protection, immunity waning, risk of reinfection, rate of recovery and other potential confounding factors, we follow the epidemiological analysis from the literature and summarize them in Supplementary Tables S1-S3. Given that the estimates for the ratio of vaccine allocation might fluctuate, we allow the parameter to vary over a continuous scale.

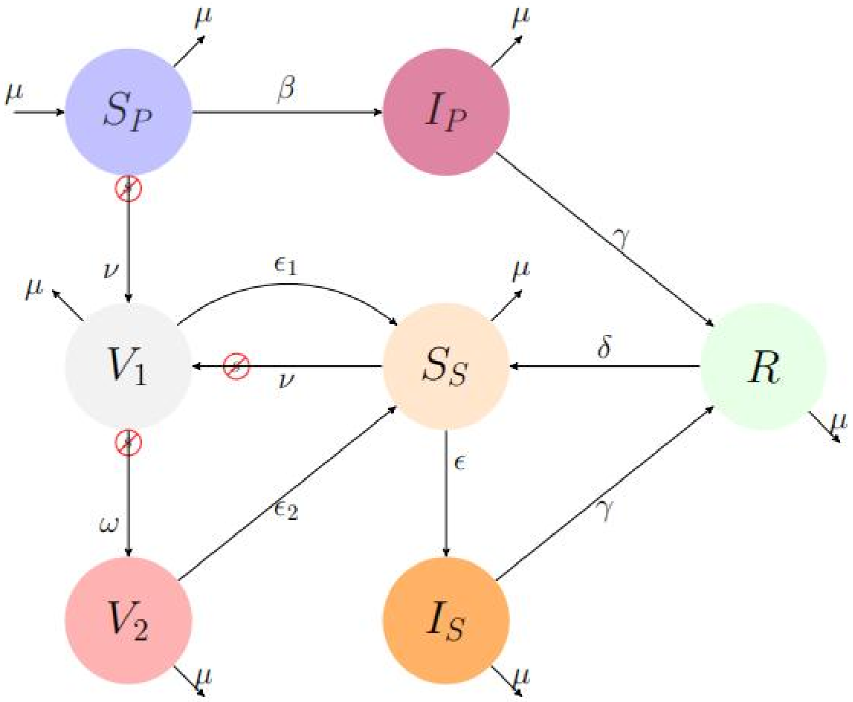


**Fig.S1: flowchart of COVID-19 transmission dynamics**

- Dose 1 vaccination
- Dose 2 vaccination
- Primary infection
- Recovered
- Partial susceptibility
- Secondary infection
- Full susceptibility

---

Suppose a susceptible-vaccinated-infected-recovered-susceptible[SVIRS] mathematical model and a population of size N (Fig. S1). The model is idential for the two settings. Assume the rate of birth is equivalent to the rate of death $\mu$, individuals recover from either primary infection or secondary infection at the rate $\gamma$, and the immunity protection wanes at the rate $\delta$. Full susceptible individuals get primary doses 1&2 vaccinated at the rate $\nu$ and $\omega$ respectively. The immunity protection of doses 1&2 declines at rate $\varepsilon_1$ and $\varepsilon_2$ respectively. $S_{vax_1}$ , $S_{vax_2}$ and denote the administration initiation timing of doses 1 and 2. $\lambda$ is the ratio of vaccines allocated from HAS to LAS. $\alpha$ is the relative infectiousness of the secondary infection in comparison with the primary infection. In the analysis, we assume this value equal to one. Hence, the total number of COVID-19 infections at equilibrium is determined by the formula (1):

$$I = I_P + \alpha I_S \qquad (1)$$

The model governing the dynamic of transmission can be expressed as the series of the following:

$$\frac{dI}{dt} = \beta I\ [S_P + \alpha S_S] - (\gamma + \mu) I \qquad (2)$$

$$\frac{dS_P}{dt} = \mu - \beta S_P I\ - (\mu + \nu) S_P \qquad (3)$$

$$\frac{dR}{dt} = \gamma I\ - (\delta + \mu) R \qquad (4)$$

$$\frac{dS_S}{dt} = \delta R + \varepsilon_1 \boldsymbol{V}_1 + \varepsilon_2 \boldsymbol{V}_2 - \ \varepsilon \beta S_S I\ - (\nu + \mu) S_S \qquad (5)$$

$$\frac{dV_1}{dt} = \nu S_P + \nu S_S - \ \varepsilon_1 V_1 - \ (\omega + \mu) V_1 \qquad (6)$$

$$\frac{dV_2}{dt} = \omega V_1 - \varepsilon_2 V_2 - \mu V_2 \quad (7)$$

If the transmission of COVID-19 reaches the equilibrium, the solution for the system is derived as follows:

$$S_P{}^* + \alpha S_S{}^* = \frac{\gamma + \mu}{\beta} \qquad (8a)$$

$$R^* = \frac{\gamma I^*}{\delta + \mu} \qquad (8b)$$

$$S_P{}^* = \frac{\mu}{\beta I^* + (\mu + \nu)} = \frac{A}{BI^* + C} \qquad (8c)$$

$$\boldsymbol{V}_1 = \frac{\nu S_P + \nu S_S}{\varepsilon_1 + \omega + \mu} \qquad (8d)$$

$$V_2 = \frac{\omega V_1}{\varepsilon_2 + \mu} \quad (8e)$$

Substituting these into the formula (5), we could obtain the following relationship:

$$\frac{\delta \gamma I^*}{\delta + \mu} + \left( \varepsilon_1 + \frac{\varepsilon_2 \omega}{\varepsilon_2 + \mu} \right) \frac{\nu S_P + \nu S_S}{\varepsilon_1 + \omega + \mu_1} - \varepsilon \beta S_S I^* - (\nu + \mu) S_S = 0 \qquad (9a)$$

$$\alpha S_S = \alpha \frac{\frac{\delta\gamma I^*}{\delta+\mu} + \left(\varepsilon_1 + \frac{\varepsilon_2\omega}{\varepsilon_2+\mu}\right)\frac{\nu S_P}{\varepsilon_1+\omega+\mu}}{\varepsilon\beta I^* + (\nu+\mu) - \left(\varepsilon_1 + \frac{\varepsilon_2\omega}{\varepsilon_2+\mu}\right)\frac{\nu}{\varepsilon_1+\omega+\mu}} = \frac{DI^* + \frac{EA}{BI^*+C}}{BI^*+F} \qquad (9b)$$

$$\frac{A}{BI^*+C} + \frac{DI^* + \frac{EA}{BI^*+C}}{BI^*+F} = Q \qquad (9c)$$

Where we assume the parameters A, B, C, D,E, F and Q satisfy the following:

$$A = \mu \qquad (10a)$$

$$B = \beta; C = \mu + \nu \qquad (10b)$$

$$D = \frac{\delta\gamma\alpha}{(\delta+\mu)\varepsilon} \qquad (10c)$$

$$Q = \frac{\gamma+\mu}{\beta} \qquad (10d)$$

$$\boldsymbol{E} = \left(\varepsilon_1 + \frac{\varepsilon_2\omega}{\varepsilon_2+\mu}\right)\frac{\nu\alpha}{(\varepsilon_1+\omega+\mu)\varepsilon} \qquad (10e)$$

$$\boldsymbol{F} = \frac{(\nu+\mu)}{\varepsilon} - \left(\varepsilon_1 + \frac{\varepsilon_2\omega}{\varepsilon_2+\mu}\right)\frac{\nu}{(\varepsilon_1+\omega+\mu)\varepsilon} \qquad (10f)$$

Hence, we can get the function of equilibrium infections expressed in (11) and (12):

$$A\left(BI^* + F\right) + DI^*\left(BI^* + C\right) + EA = Q\left(BI^* + C\right)\left(BI^* + F\right) \qquad (11)$$

$$f\left(I^*\right) = \left(QB^2 - DB\right)I^{*2} + \left(QBF + QBC - DC - AB\right)I^* + \left(QCF - EA - AF\right) \quad (12)$$

We additionally assume:

$$a = QB^2 - DB; \quad b = QBF + QBC - DC - AB; \quad c = QCF - EA - AF$$

The solution at equilibrium satisfies formula (13):

$$I^* = \frac{-b + \sqrt{b^2 - 4ac}}{2a} \qquad (13)$$

We then consider the scenario where ratio $\lambda$ of of the population are moving across the two settings, for each setting $i, j\,(i, j = 1,2)$ we can derive:

$$I_T = (1-\lambda)I_H + \lambda I_L \qquad (14a)$$

$$\frac{dS_{P,i}}{dt} = \mu - \beta S_{P,i}\left[(1-\lambda)I_{T,i} + \lambda I_{T,j}\right] - \left(s_{vax1}v_i + \mu\right)S_{P,i} \qquad (14b)$$

$$\frac{dI_{P,i}}{dt} = \beta S_{P,i}\left[(1-\lambda)I_{T,i} + \lambda I_{T,j}\right] - (\gamma + \mu)I_{P,i} \qquad (14c)$$

$$\frac{dR_i}{dt} = \gamma\left[(1-\lambda)I_{T,i} + \lambda I_{T,j}\right] - (\delta + \mu)R_i \qquad (14d)$$

$$\frac{dS_{S,i}}{dt} = \delta R_i + \varepsilon_1 V_{1,i} + \varepsilon_2 V_{2,i} - \varepsilon \beta S_{S,i}\left[(1-\lambda) I_{T,i} + \lambda I_{T,j}\right] - \left(s_{vax1} \nu_i + \mu\right) S_{S,i} \qquad (14e)$$

$$\frac{dI_{S,i}}{dt} = \varepsilon \beta S_{S,i}\left[(1-\lambda) I_{T,i} + \lambda I_{T,j}\right] - (\gamma + \mu) I_{S,i} \qquad (14f)$$

$$\frac{dV_{1,i}}{dt} = s_{vax1} \nu_i S_{P,i} + s_{vax3} \nu_i S_{S,i} - \varepsilon_1 V_{1,i} - \left(s_{vax2} \omega_i + \mu\right) V_{1,i} \qquad (14g)$$

$$\frac{dV_{2,i}}{dt} = s_{vax2} \omega_i V_{1,i} - \varepsilon_2 V_{2,i} - \mu V_{2,i} \qquad (14h)$$

$$S_{vax1} = \begin{cases} 0, & t < t_{vax1} \\ 1, & t \geq t_{vax1} \end{cases} \qquad (15)$$

$$S_{vax2} = \begin{cases} 0, & t < t_{vax2} \\ 1, & t \geq t_{vax2} \end{cases} \qquad (16)$$

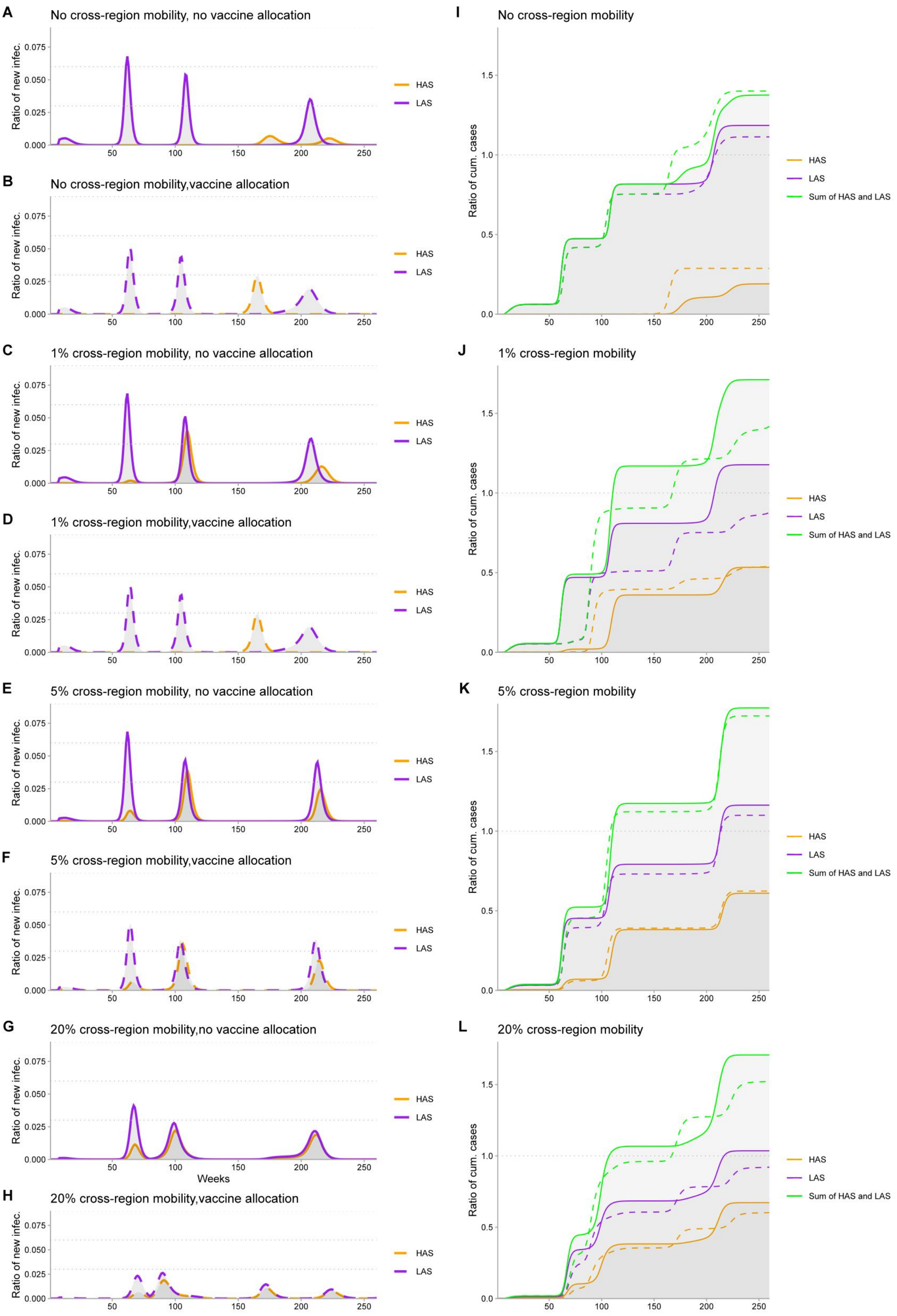


Fig.S2 Impact of vaccine allocation and population mobility on the transmission dynamics of COVID-19 pandemic. Allocation ratio of vaccines from HAS to LAS is 0.1.

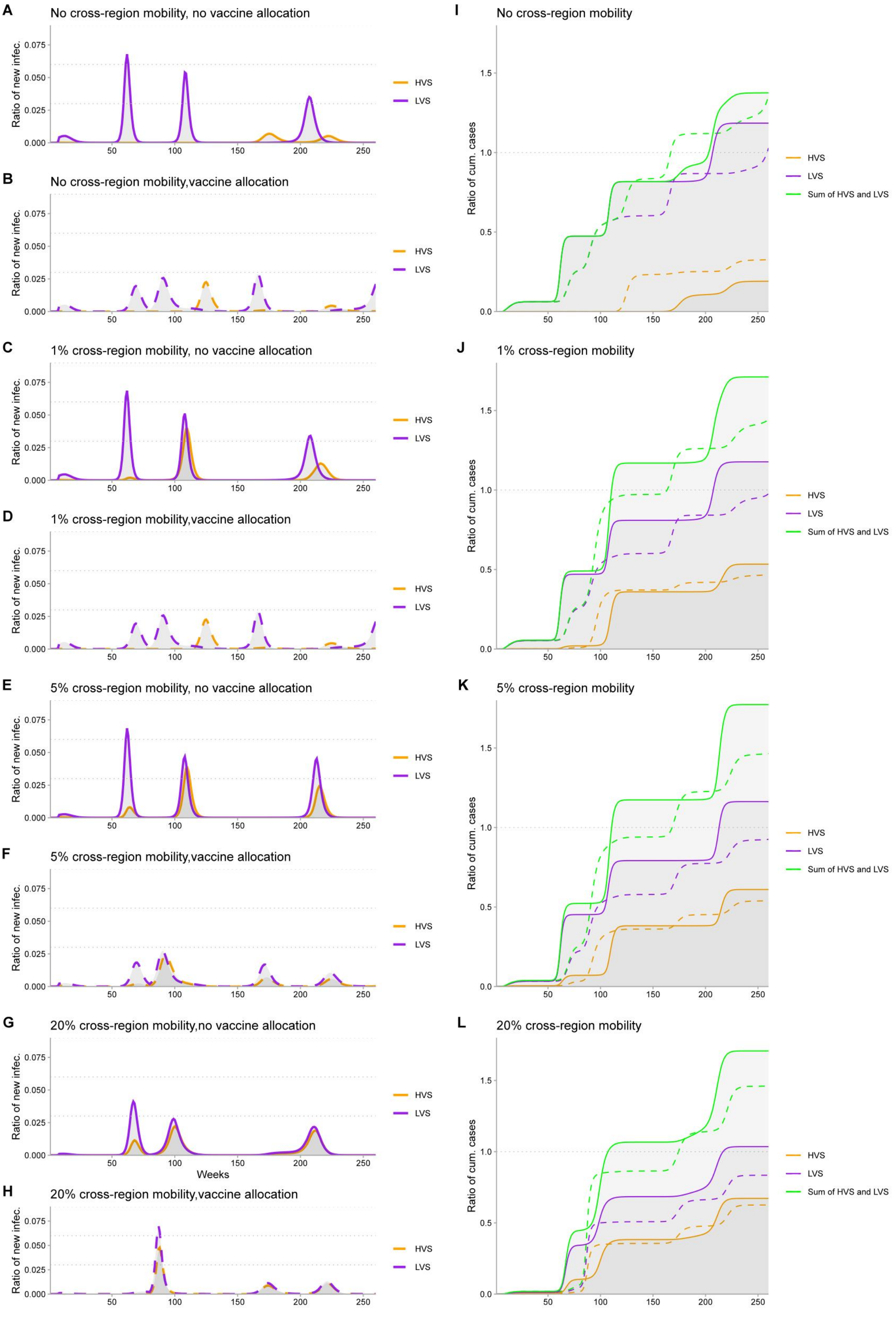


Fig.S3 Impact of vaccine allocation and population mobility on the transmission dynamics of COVID-19 pandemic. Allocation ratio of vaccines from HAS to LAS is 0.3.

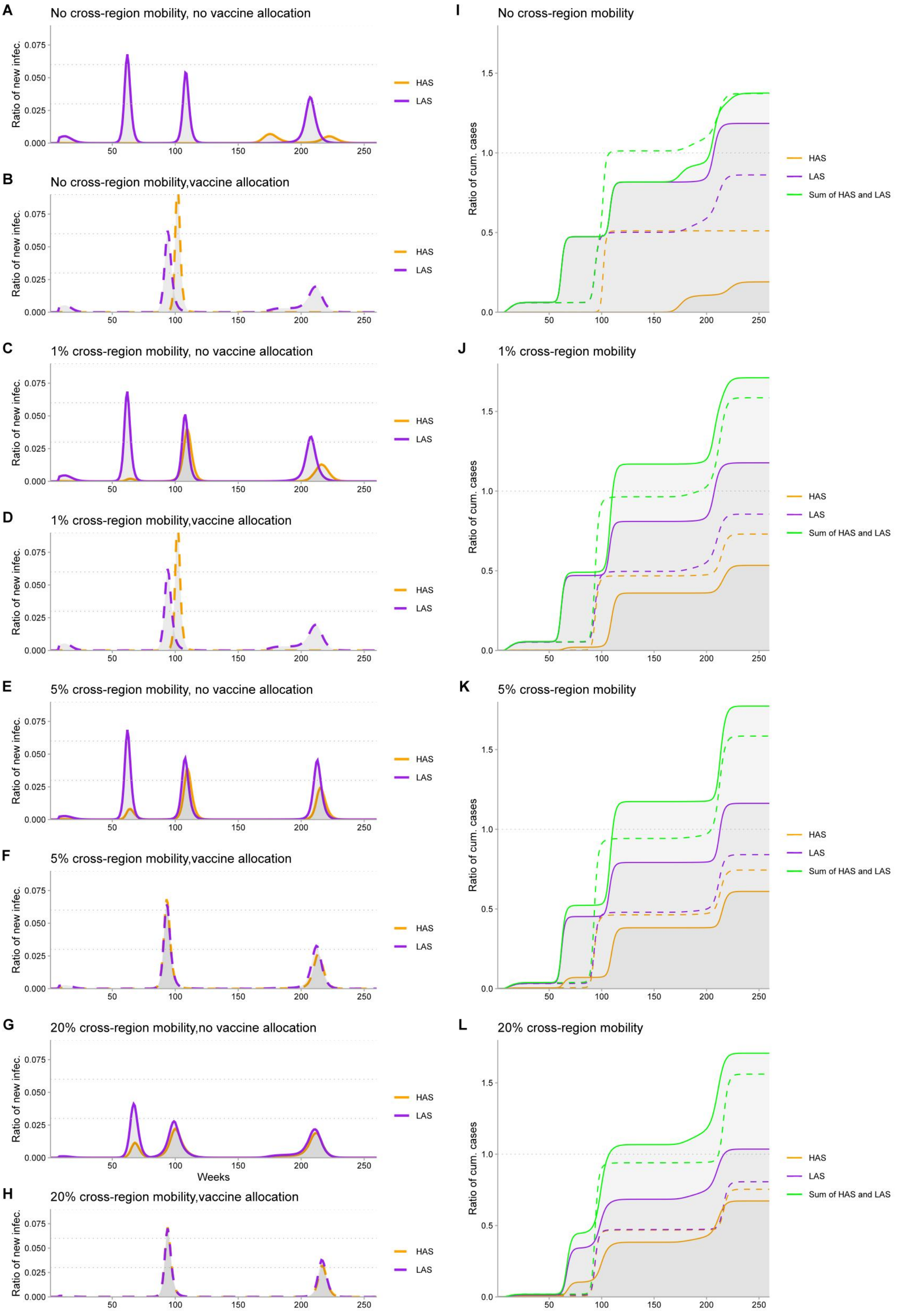


Fig.S4 Impact of vaccine allocation and population mobility on the transmission dynamics of COVID-19 pandemic.
Allocation ratio of vaccines from HAS to LAS is 0.7.

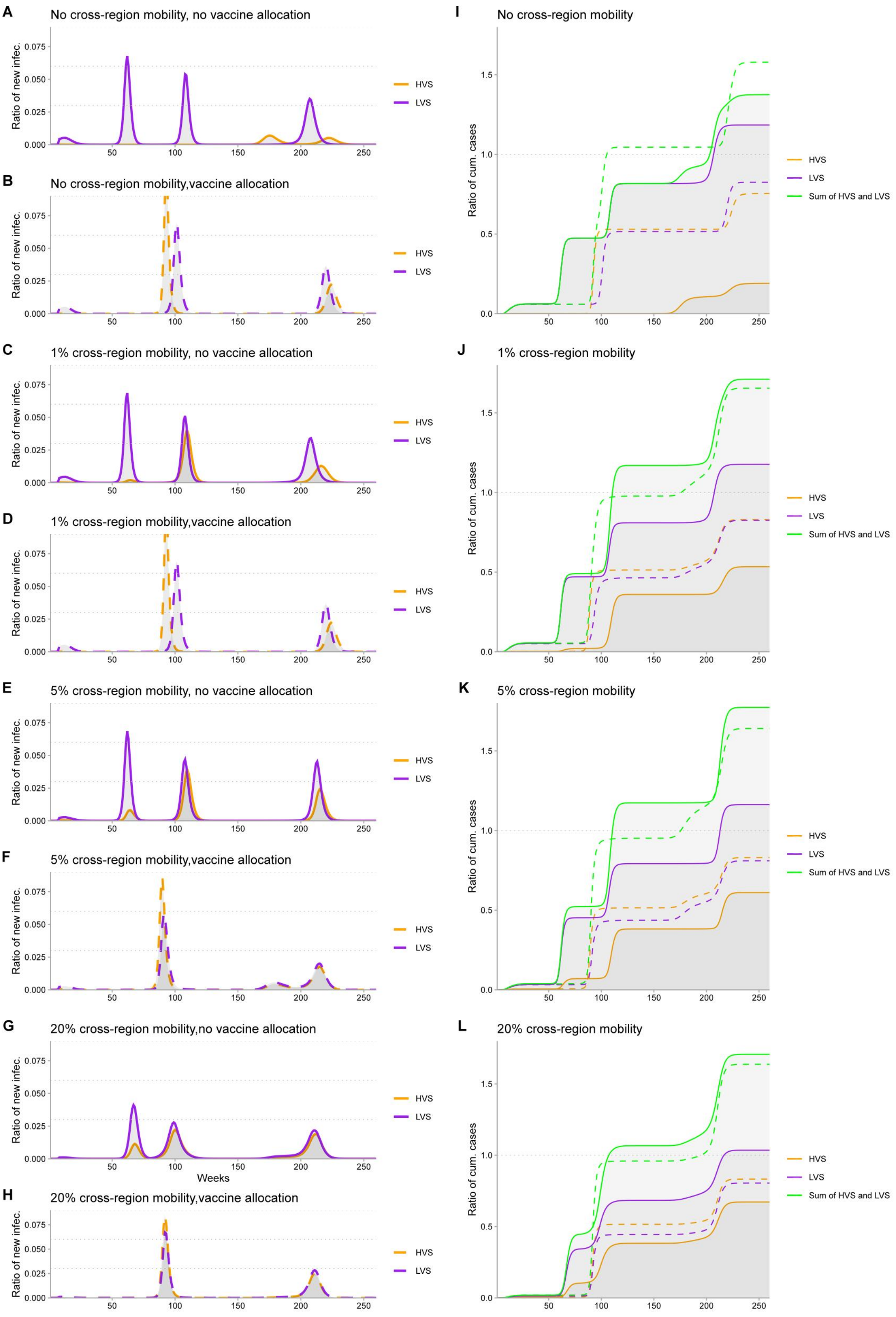


Fig.S5 Impact of vaccine allocation and population mobility on the transmission dynamics of COVID-19 pandemic.
Allocation ratio of vaccines from HAS to LAS is 0.9.

**Table S1 Simulation parameters for Fig. 2 and FigS2-S5**

| Name | Description | Baseline Value | References |
|---|---|---|---|
| $\alpha$ | relative infectiousness of $I_S$ | 1 | [2][3][4][5] |
| $\beta$ | transmission rate | $R*\gamma$ | [2][3] |
| $t_{switch}$ | susceptibility after the waning of dose 1 immunity | 16 week | Assumed |
| $\varepsilon$ | rate of secondary infection | 0.55 per week | Assumed |
| $\mu$ | birth rate or death rate | 0.02 per week | [2][3][4][5] |
| $\gamma$ | the recovery rate of COVID-19-positive patients | 1.4 | [2][3][4][5] |
| $v$ | vaccination rate of dose 1 | 0.02 per week | [2][3][4][5] |
| $\omega$ | vaccination rate of dose 2 | HAS:0.03 $\rightarrow$0.02 per week<br>LAS:0.01 $\rightarrow$0.02 per week | Assumed |
| $\delta$ | waning rate to secondary susceptibility | 1 per week | Assumed |
| $\{S_{vax_1}, S_{vax_2}\}$ | initiation of dose 1&2 | {16,16}[week] | Assumed |
| $\mathbf{S}_P$ | initial size of the full susceptible population | 1- $I_0$ | [2][3][4][5] |
| $N$ | size of population | 1 | Assumed |
| $I_0$ | initial size of infection | 1e-9 | [2][3][4][5] |
| $R$ | reproduction number | 2.3 | [2][3][4][5] |
| $\lambda$ | rate of infected population mobility | {0,0.01,0.05,0.2} per week | Assumed |
| $\rho$ | ratio of allocation | {0.5(Fig. 2), 0.1(Fig.S2), 0.3(Fig.S3),0.7(Fig.S4),0.9 (Fig.S5)} | Assumed |

## Table S2    Simulation parameters for Fig. 3

| Name | Description | Baseline Value | References |
|---|---|---|---|
| $n$ | length of observation | 260 weeks | Assumed |
| $\varepsilon$ | rate of secondary infection | {0.5(normal), 0.6(medium), 0.7(high)} per week | Assumed |
| $\varepsilon_1$ | waning rate of immunity protection of dose 1 | 0.5 per day | Assumed |
| $\varepsilon_2$ | waning rate of immunity protection of dose 1 | 1 per day | Assumed |
| $\gamma$ | the recovery rate of COVID-19-positive patients | 0.2 | [2][3][4][5] |
| $\nu$ | vaccination rate of dose 1 | {0.0001,0.003,0.005} per week | Assumed |
| $\frac{\omega_H}{\omega_L}$ | rate ratio of HAS to LAS for dose 2 | $\frac{\omega_H}{\omega_L}=1;\frac{\omega_H}{\omega_L}=\frac{1}{3};\frac{\omega_H}{\omega_L}=3$ | Assumed |
| $\omega$ | rate of dose 2 | {0.2(normal),0.5(speed-enhanced)} year | Assumed |
| $\beta$ | rate of transmission | {2.4/5(Baseline), 1.2*2.4/5(Mild Variant), 2.4*2.4/5(Severe Variant),}[week] | Assumed |

### Table S3 Simulation parameters for Fig. 4

| Name | Description | Baseline Value | References |
|---|---|---|---|
| $\alpha$ | relative infectiousness of $I_S$ | 1 | Assumed |
| $\beta$ | transmission rate | $R*\gamma$ | Assumed |
| $t_{switch}$ | susceptibility after the waning of dose 1 immunity | 16 week | Assumed |
| $\varepsilon$ | rate of secondary infection | {0.2, 0.5, 0.7} per week | Assumed |
| $\mu$ | birth rate or death rate | 0.02 per week | [2][3][4][5] |
| $\gamma$ | the recovery rate of COVID-19-positive patients | 1.4 | [2][3][4][5] |
| $\nu$ | vaccination rate of dose 1 | 0.003 per week | Assumed |
| $\omega$ | rate ratio of HAS to LAS for dose 2 | $\frac{\omega_H}{\omega_L}=1; \frac{\omega_H}{\omega_L}=\frac{1}{3}; \frac{\omega_H}{\omega_L}=3$ | Assumed |
| $\delta$ | rate of waning to secondary susceptibility | {0.5,1,1.5} year | Assumed |
| $\{S_{vax_1}, S_{vax_2}\}$ | initiation of dose 1&2 | {16,16}[week] | Assumed |
